# Characterizing Patient Representations for Computational Phenotyping

**DOI:** 10.1101/2022.07.26.22278073

**Authors:** Tiffany J. Callahan, Adrianne L. Stefanksi, Danielle M. Ostendorf, Jordan M. Wyrwa, Sara J. Deakyne Davies, George Hripcsak, Lawrence E. Hunter, Michael G. Kahn

## Abstract

Patient representation learning methods create rich representations of complex data and have potential to further advance the development of computational phenotypes (CP). Currently, these methods are either applied to small predefined concept sets or all available patient data, limiting the potential for novel discovery and reducing the explainability of the resulting representations. We report on an extensive, data-driven characterization of the utility of patient representation learning methods for the purpose of CP development or automatization. We conducted ablation studies to examine the impact of patient representations, built using data from different combinations of data types and sampling windows on rare disease classification. We demonstrated that the data type and sampling window directly impact classification and clustering performance, and these results differ by rare disease group. Our results, although preliminary, exemplify the importance of and need for data-driven characterization in patient representation-based CP development pipelines.

## Introduction

Clinical phenotyping, or the classification of patients by disease status, is a technique designed to provide clinicians and clinical researchers with important insights into the development, progression, and outcome of disease(s). Computational phenotyping is the process of applying computer-executable algorithms to clinical data in order to derive cohorts or groups of patients with and without a specific disease or phenotype of interest. Traditionally, computational phenotypes (CP) have largely been expert-defined and leveraged electronic health record (EHR) data. While these methods are likely to produce highly accurate cohorts, they require significant development resources, rely on a wide variety of domain experts, and are limited to known phenotypes and well-characterized diseases^1–3^.

Representation learning techniques, which meaningfully transform a wide range of complex heterogeneous data into representations well-suited for downstream learning^4^, are a promising approach for improving CP development without sacrificing valuable information, data, or the potential for novel discovery. As noted in a recent systematic review, the use of these methods to learn patient representations from EHR data has nearly tripled since 2015^5^, which speaks to the utility of these methods to solve a wide variety of problems within the clinical domain. Although there are many methods for deriving patient representations^5^, vector-based methods, such as variational autoencoders or word embeddings are the most frequently used techniques for deriving patient representations. These techniques have been used to learn static and temporal representations of patients and clinical concepts^6^, jointly represent temporal and atemporal patient information^7,8^, and incorporate both structured and unstructured data^9^.

Despite successful implementations of these methods, their adoption and use in clinical settings remains largely unrealized, possibly due to two primary and related challenges^10–12^. The first challenge is *explainability*. The resulting patient representations usually consist of large feature matrices or one-dimensional arrays of continuous numbers, which can be difficult or even impossible to interpret without additional context or metadata. Further, models learned from patient representations methods can be difficult to explain without including external context or metadata. The second challenge is *scalability*. The majority of existing applications tend to utilize either small expert-selected code sets or all available data when learning patient representations. Leveraging expert-derived code sets may result in more accurate representations at the cost of novel discovery^13,14^. In contrast, learning representations from all available patient information may present opportunities for discovery and decrease development time, at the cost of creating “noisier” representations, which may be more difficult to interpret.

Unlike other fields which perform comprehensive diagnostics or characterization prior to analysis, the evaluation of patient representation learning methods has largely been limited to the context of a specific downstream use case and is usually performed as part of model interpretation. While it cannot solve all of the aforementioned challenges, data-driven characterization of patient representations, independent of model development, may provide invaluable and unexpected insights and is an important first step towards understanding whether these methods can be used to help automate CP development. To this end, we sought to answer the following questions:

1. What combinations of data type and sampling window create the best patient representations and does performance differ by disease group?
2. How does data-driven characterization of patient representation impact the explainability of downstream tasks like clustering?

To address these questions, we examined the impact of sampling window (i.e., all data, all data before the index date, and all data available after the index date), and data type (i.e., only conditions, only drug exposures, only measurements, and all data types) on patient representations within the context of rare disease. The work presented in this paper makes the following contributions:

- We performed, to the best of our knowledge, the first comprehensive characterization of patient representations learned from EHR data.
- As an evaluation, the similarity between any two patients was used as a diagnostic biomarker and assessed by its effectiveness for identifying similar patients, which is generalizable to a wide-range of patient representation learning algorithms and downstream learning tasks.
- We conducted extensive ablation studies to examine the impact of learning patient representations using different data types and sampling windows. We performed these studies on four different types of rare disease patients and medically complex controls. We evaluated the resulting representations using two tasks: (i) Leave-one-patient-out classification, which was assessed using a wide variety of performance metrics and plots; and (ii) K-means clustering, which included domain expert review of the most important features for each cluster.

An extended version of this work, which includes the examination of additional data types and more extensive analytics, can be found here: https://doi.org/10.5281/zenodo.5716401 (see Section 3.1.2).

## Methods

### Clinical Data

A subset of de-identified data was extracted in April 2018 from Children’s Hospital Colorado (CHCO) and deposited in the Health Data Compass Warehouse at the University of Colorado Anschutz Medical Campus. Data conformed to the structure defined by the PEDSnet Observational Medical Outcomes Partnership (OMOP) common data model (CDM) version 2.8, which is an adaptation of the OMOP CDM version 5.0^15,16^. Use of these data was approved by the Colorado Multiple Institutional Review Board (15-0445).

### Patient Cohorts

Two groups of pediatric patients were utilized for the current project. Cases included rare disease patients with at least 10 visits and a primary diagnosis of congenital hypothyroidism (CAH), cystic fibrosis (CF), phenylketonuria (PKU), or sickle cell disease (SCD). Control patients were also required to have at least 10 visits and no occurrence of the diagnosis codes used to identify the rare disease patients. The SQL queries used to identify rare disease and control patients are publicly available as a GitHub Gist (https://gist.github.com/callahantiff/b72339a8d6fb7de31e1912039947605a).

### Patient Representations

Patient representations were learned using a bag-of-words (BoW)^17^ vector space model with term-frequency inverse-document frequency (TF-IDF)^18^ and L2 normalization. While more complex methods exist, this approach was chosen because it has been proven to be robust for a wide range of biomedical tasks^19–22^ and because it is one of the more transparent and explainable vector-based patient representation learning methods. All diagnosis codes used to identify rare disease patients were excluded from representations (see SQL query for concept sets).

### Evaluation

#### Classification

Ablation studies were performed to examine the impact of altering *data type* (i.e., only conditions, only drug exposures, only measurements versus “all data types’’ defined as the combination of all three prior data types) and *sampling window* (i.e., all available data, all data available before the index date, and all data available after the index date). For control patients, a single sampling window that included all available concepts was used. The resulting representations were assessed by how well they were able to accurately identify similar patients of a specific disease type. For this task, the similarity between any two patients was used as a diagnostic biomarker and assessed by its effectiveness for identifying similar patients. The Youden Index or *J* statistic^23^, is a receiver operating curve (ROC) summary statistic that takes a continuous variable as input and outputs a diagnostic threshold that can be used to determine which patients are and are not “similar enough” to a query patient. This approach was applied for each patient, and the diagnostic effectiveness of different patient representations was assessed using leave-one-patient-out classification, where each rare disease patient was classified against all other patients. The following metrics used included: specificity, recall, precision, accuracy, F_1_ Score, and area under the ROC curve (AUC). Metrics were reported as the average score for each rare disease across 10 iterations. For each iteration, the diagnostic effectiveness of a specific patient representation-derived configuration, for each rare disease, was evaluated on its ability to identify patients of the same disease type from a pool of 1,000 randomly sampled control patients and patients from all other rare disease groups.

#### Clustering

The explainability of the patient representations were assessed using K-Means^24^ with a predefined *k* of four when only assessing rare disease patients and five when assessing rare disease and control patients. The resulting clusters were evaluated using the Adjusted Rand Index (ARI)^25^, Adjusted Mutual Information (AMI)^26^, and Cluster Purity (Purity)^27^. All metrics were scored 0-1, where 1.0 indicates that the true and predicted clustering assignments were identical. In addition to these metrics, two ways of measuring the homogeneity of a cluster were also examined: (i) <u>*Cluster Breakdown*</u>: % of rare disease patients in each rare disease group out of the total patients in that cluster; (ii) <u>*Disease Breakdown*</u>: % of rare disease patients for each disease group out of the total number of patients in each rare disease group in that cluster. Taken together, these metrics provide a means for interpreting the distribution of a cluster with respect to each disease (e.g., 73% of the patients in Cluster 1 had CF [cluster breakdown], which represents 45% of the total sample of CF patients [disease breakdown]). Finally, the clustering results for the best performing representations were further examined by investigating the three most frequent features and the three most frequent features that were unique to each cluster. This output was reviewed with domain experts to assess the clinical relevance of the resulting clusters.

All preprocessing of data, construction of patient representations, and evaluation tasks were performed using Python 3.6.2 on a machine running macOS Sierra with 16 GB of RAM and a 2.7 GHz processor with 8 cores. The Python scikit-learn (v.0.22.1) library was used to create the BoW+TF-IDF representations and perform clustering and ROC plots were visualized using Matplotlib (v.3.3.2). All code is publicly available as a GitHub Gist: https://gist.github.com/callahantiff/1b6b0530bd057527108ef335aea96a97.

## Results

A total of 2,643 rare disease patients (CAH: *n*=760, CF: *n*=835, PKU: *n*=235, SCD: *n*=813) and 10,000 control patients were obtained. On average, there were slightly more female (52%) than male patients with an average age at first recorded relevant diagnosis (index date) of 5.83 years (CAH: *M*=3.23; CF: *M*=6.51; PKU: *M*=11.10; and SCD: *M*=5.35; Control: *M*=5.85). Note that age was calculated from the earliest occurrence of a relevant diagnosis code in each patient’s record. For rare disease patients, the earliest occurrence of a relevant diagnosis code was used as the index date (see SQL query above for concept sets). For control patients, age was calculated from the earliest occurrence of the most frequently occurring diagnosis code. Table 1 contains the counts of patients and unique concepts organized by data type and sampling window. The number of features available for each representation ranged from as few as 271 to over 9,000 concepts.

**Table 1.** Counts of Patients and Unique Concepts by Disease Group and Representation.

| Data Type | Unique<br>Concept<br>Count | Disease Cohort |  |  |  |  | Patient<br>Count |
| --- | --- | --- | --- | --- | --- | --- | --- |
|  |  | CF | CAH | PKU | SCD | Control |  |
| All Concepts |  |  |  |  |  |  |  |
| Conditions | 6493 | 828 | 696 | 173 | 813 | 10000 | 12510 |
| Drug Exposure | 2310 | 820 | 750 | 91 | 805 | 8656 | 11122 |
| Measurement | 271 | 833 | 760 | 234 | 814 | 9704 | 12345 |
| All Data Types | 9074 | 835 | 760 | 235 | 814 | 10000 | 12644 |
| Concepts Before Index Date |  |  |  |  |  |  |  |
| Conditions | 5796 | 530 | 497 | 86 | 705 | 10000 | 11818 |
| Drug Exposure | 1774 | 455 | 535 | 11 | 535 | 8656 | 10192 |
| Measurements | 263 | 699 | 729 | 198 | 741 | 9704 | 12071 |
| All Data Types | 7833 | 747 | 739 | 201 | 752 | 10000 | 12439 |
| Concepts After Index Date |  |  |  |  |  |  |  |
| Conditions | 6377 | 828 | 683 | 172 | 811 | 10000 | 12494 |
| Drug Exposure | 2253 | 813 | 732 | 90 | 780 | 8656 | 11071 |
| Measurements | 266 | 832 | 759 | 234 | 811 | 9704 | 12340 |
| All Data Types | 8896 | 835 | 759 | 235 | 812 | 10000 | 12641 |

Acronyms - CAH: Congenital Hypothyroidism; CF: Cystic Fibrosis; PKU: Phenylketonuria; and SCD: Sickle Cell Disease.

### Classification

Table 2 contains the classification results. To aid in interpreting the results, highlighting was used to point out the best (green) and worst (red) performance for each metric within each disease, data type, and sampling window.

**Table 2.**
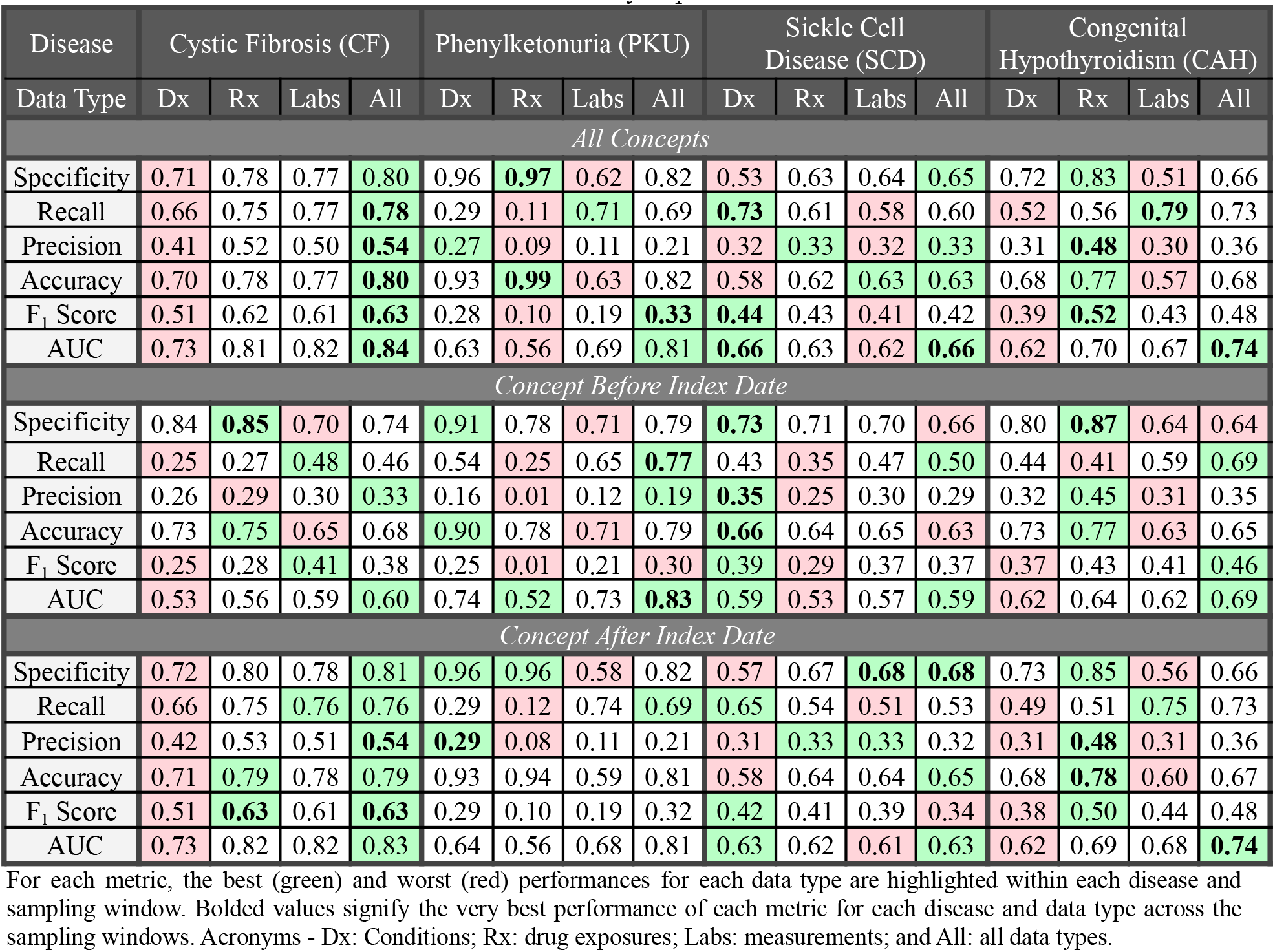
Rare Disease and Control Patient Performance by Representation.

The single best performance for each metric is bolded for each disease and data type across all sampling windows. Results are reported by disease for three different views: data type, sampling window, and data type by sampling window. The best performance was determined by taking the highest average score across all metrics. For brevity, we report AUC for all views. For the sampling window and data type views, we report the mean and range. For the sampling window by data type view, we report the AUC for each sampling window.

#### Data Type

The ROC plots in Figure 1 visualizes the performance of representations learned using all concepts for each data type by rare disease group. Using concepts from all data types resulted in the best performance for **CF** (*AUC*=0.76, 0.60-0.84) and **PKU** patients (*AUC*=0.82, 0.81-0.83). Using only drug exposure concepts worked best for **CAH** patients (*AUC*=0.68, 0.64-0.62) and using only condition concepts worked best for **SCD** patients (*AUC*=0.63, 0.59-0.66). Using only condition concepts resulted in the worst performance for **CF** (*AUC*=0.66, 0.53-0.73) and **CAH** patients (*AUC*=0.62, 0.62-0.62). Finally, using only drug exposure concepts resulted in the worst performance for **PKU** (*AUC*=0.55, 0.52-0.56) and **SCD** patients (*AUC*=0.59, 0.53-0.63).

**Figure 1.**
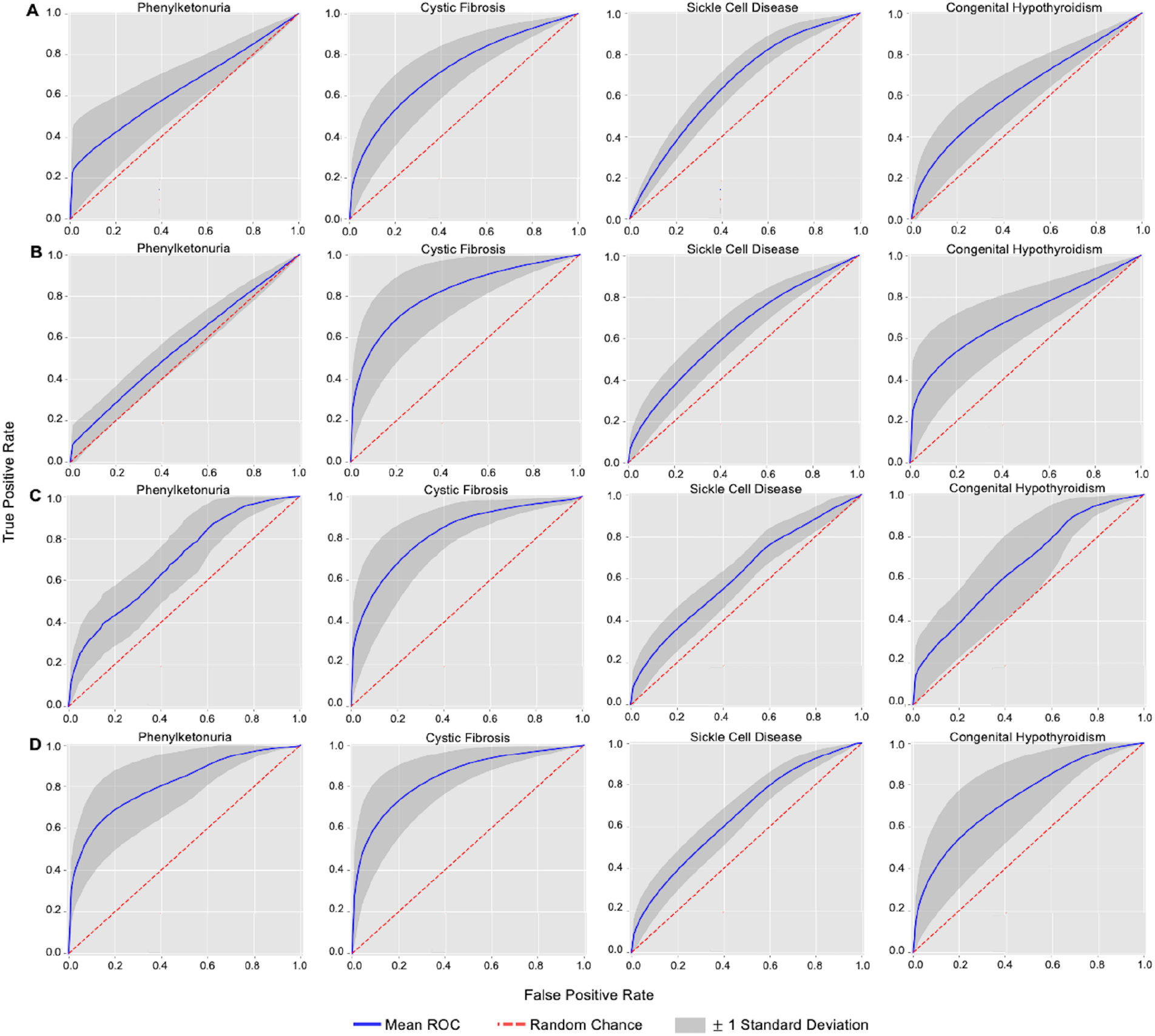
ROC plots for rare disease patient representations learned using all available (A) condition concepts, (B) drug exposure concepts, (C) measurement concepts, and (D) concepts from all data types.

#### Sampling Window

Using concepts after the index date resulted in the best performance for **CF** (*AUC*=0.80, 0.73-0.83) and **CAH** (*AUC*=0.68, 0.62-0.74) patients. Using all concepts resulted in the best performance for **PKU** (*AUC*=0.67, 0.56-0.81) and **SCD** (*AUC*=0.64, 0.62-0.66) patients. For all rare disease groups, the worst performing patient representations were built using concepts before the index date (CF: *AUC*=0.57, 0.53-0.60; PKU: *AUC*=0.51, 0.01-0.95; CAH: *AUC*=0.64, 0.62-0.69; SCD: *AUC*=0.57, 0.53-0.59).

#### Data Type by Sampling Window

Using all data types at each sampling window resulted in the best performance for **CF** (All: *AUC*=0.84, Before: *AUC*=0.60; After: *AUC*=0.83) and **PKU** (All: *AUC*=0.81; Before: *AUC*=0.83; After: *AUC*=0.81) patients. While using all data type concepts also resulted in the best performance for **SCD** patients (Best: 0.48; *AUC*=0.66), the best performance before the index date was found when using only condition concepts (*AUC*=0.59), and when using only those concepts after the index date, the best performance was found when using only drug exposure concepts (*AUC*=0.62). Finally, for **CAH** patients, the best performance was achieved for all sampling windows when using drug exposure concepts (All: *AUC*=0.70, Before: *AUC*=0.64; After: *AUC*=0.69).

### Clustering

#### Clustering Performance

Table 3 contains the clustering results using the same highlighting and bolding strategy that was applied in Table 2. Results are presented in two waves: only rare disease patients vs rare disease and control patients combined. The best performance by sampling window and data type, the mean and range are presented for each clustering metric. When examining the performance of each data type by sampling window, a single result is presented for each metric.

**Table 3.** Rare Disease Patient Clustering Metrics by Data Type and Sampling Window.

| Metric | Rare Disease Patients |  |  |  | Rare Disease and Control Patients |  |  |  |
| --- | --- | --- | --- | --- | --- | --- | --- | --- |
|  | Dx | Rx | Labs | All | Dx | Rx | Labs | All |
| *All Concepts* |  |  |  |  |  |  |  |  |
| Adjusted Rand Index | **0.288** | **0.416** | 0.211 | **0.347** | 0.038 | 0.035 | 0.018 | 0.033 |
| Adjusted Mutual Information | **0.262** | 0.421 | 0.208 | **0.318** | 0.071 | 0.096 | 0.065 | 0.078 |
| Purity | 0.662 | 0.752 | 0.563 | 0.669 | 0.799 | 0.778 | 0.786 | 0.791 |
| *Concepts Before Index Date* |  |  |  |  |  |  |  |  |
| Adjusted Rand Index | 0.211 | 0.253 | 0.057 | 0.121 | 0.015 | 0.021 | 0.013 | 0.021 |
| Adjusted Mutual Information | 0.201 | 0.241 | 0.067 | 0.126 | 0.041 | 0.034 | 0.028 | 0.047 |
| Purity | 0.634 | 0.665 | 0.407 | 0.517 | **0.846** | **0.849** | **0.804** | **0.804** |
| *Concepts After Index Date* |  |  |  |  |  |  |  |  |
| Adjusted Rand Index | 0.279 | 0.413 | **0.236** | 0.314 | 0.021 | 0.040 | 0.010 | 0.046 |
| Adjusted Mutual Information | 0.246 | **0.428** | **0.230** | 0.292 | 0.063 | 0.092 | 0.039 | 0.093 |
| Purity | 0.660 | 0.748 | 0.595 | 0.640 | 0.800 | 0.782 | 0.786 | 0.791 |

For each metric, the best (green) and worst (red) performances for each data type are highlighted within each group and sampling window. Bolded values signify the very best performance of each metric for each data type across the sampling windows. Acronyms - Dx: Conditions; Rx: drug exposures; Labs: measurements; and All: all data types.

##### Rare Disease Patients Only

<u>Sampling Window.</u> The best performing sampling window was found when using all concepts (ARI=0.32, 0.21-0.42; AMI=0.30, 0.21-0.42; Purity=0.66, 0.56-0.75) and the worst performance was found for concepts before the index date (ARI=0.16, 0.06-0.25; AMI=0.16, 0.07-0.24; Purity=0.57, 0.41-0.67). <u>Data Type</u>. When examining the results by data type, the best performance was found when using only drug exposure concepts (ARI=0.36, 0.25-0.42 AMI=0.36, 0.24-0.43; Purity=0.72, 0.67-0.75) and the worst performance was achieved when using only measurement concepts (ARI=0.17, 0.06-0.24; AMI=0.17, 0.07-0.23; Purity=0.52, 0.41-0.60). <u>Data Type by Sampling Window</u>. Regardless of sampling window, the best performance was found when using only drug exposure concepts (All: ARI=0.42, AMI=0.42, Purity=0.75; Before: ARI=0.25, AMI=0.24, Purity=0.67; After: ARI=0.41, AMI=0.43, Purity=0.75).

##### Rare Disease and Control Patients Combined

<u>*Sampling Window*</u>. The best performing sampling window was found when using all concepts (*ARI*=0.03, 0.02-0.04; AMI=0.08, 0.07-0.10; Purity=0.79, 0.78-0.80) and the worst performance was found for concepts before the index (*ARI* =0.02, 0.01-0.02; AMI=0.04, 0.03-0.05; Purity=0.83, 0.80-0.85). <u>*Data Type*</u>. When examining the results by data type, the best performance was found for all data type concepts (*ARI*=0.03, 0.02-0.05; AMI=0.07, 0.05-0.09; Purity=0.80, 0.79-0.80) and the worst performance was found when using only measurement concepts (*ARI*=0.01, 0.01-0.02; AMI=0.04, 0.03-0.07; Purity=0.80, 0.79-0.80). <u>*Data Type by Sampling Window*</u>. When using all concepts, the best performance was found when using only condition concepts (*ARI*=0.04; *AMI*=0.07; *Purity*=0.80). When using only those concepts before the index, the best performance was found when using only drug exposure concepts (*ARI*=0.02; *AMI*=0.03; *Purity*=0.85). Finally, when using only those concepts after the index, the best performance was found when using all data type concepts (*ARI*=0.05; *AMI*=0.09; *Purity*=0.80).

#### Important Cluster Features

Additional statistics are provided for the single best performing data type and sampling window found for rare disease patients only (i.e., all data types concepts after the index) and rare disease and control patients combined (i.e., all drug exposure concepts) clusters. For each cluster, the patient breakdown by cluster and disease are reported. We also report the top-three most frequent concepts and the top-three most frequent unique concepts for each cluster which includes the OMOP concept identifier and its frequency. To shorten the label of drug exposure concepts, the brand names were used, unless multiple formulations of the same drug appeared in the same cluster.

Rare Disease Patients Only - All Available Drug Exposure Concepts

- **Cluster 1**. Contained 1,184 concepts (313 unique) and 643 patients who were 99.7% CF (78.2% of all CF patients). *Top-three concepts*: dornase alfa (19076733, n=84) and two formulations of Creon (40166160, n=80 and 40166176, n=74). *Top-three unique concepts*: Creon (40166160; n=80) and two formulations of Zenpep (40164021; n=58 and 40164029; n=55).
- **Cluster 2**. Contained 1,327 concepts (437 unique) and 745 patients who were 65.5% SCD (60.6% of all SCD patients) and 26.9% CAH (26.7% of all CAH patients). *Top-three concepts*: RotaTeq (19130404; n=139), Prevnar 13 (40173508; n=134), and varicella-zoster virus vaccine (46275140; n=114). *Top-three unique concepts*: Haemophilus influenzae b vaccine (46276401; n=62), bordetella pertussis vaccine (42800271; n=49), and Rotarix (19131940; n=44).
- **Cluster 3**. Contained 411 concepts (25 unique) and 471 patients who were 92.8% CAH (58.3% of all CAH patients). *Top-three concepts*: three strengths of levothyroxine sodium (40174880; n=209, 40175174; n=189, and 40174907; n=170). *Top-three unique concepts*: two strengths of liothyronine sodium 40174292; n=3 and 40174879; n=2) and Moxeza (402320402; n=2).
- **Cluster 4**. Contained 3,438 concepts (331 unique) and 2,051 patients who were 47.6% SCD (35.9% of all SCD patients), 21.6% CF (16.0% of all CF patients), 18.5% CAH (14.9% of all CAH patients), and 12.4% PKU (82.4% of all PKU patients). *Top-three concepts*: ibuprofen (19019050; n=102), amoxicillin (19073186; n=80), and albuterol (42903088; n=54). *Top-three unique concepts*: Kuvan tablets and powder (44785377; n=1 and 40239923; n=16), and NuvaRing (43012433; n=7).

Rare Disease and Control Patients Combined - All Data Type Concepts After Index

- **Cluster 1**. Contained 3,497 concepts (250 unique) and 2,323 patients who were 72.1% control (16.7% of control patients), 17.1% CAH (52.3% of all CAH patients), and 4.8% PKU (47.2% of all PKU patients). *Top-three concepts*: body height (3023540; n=670), systolic blood pressure (3004249; n=458), and diastolic blood pressure (3012888; n=456). *Top-three unique concepts*: two strengths of isotretinoin 30 mg (19102526; n=26 and 984237; n=17) and Sodium [Moles/volume] in Serum or Plasma (3019550; n=7).
- **Cluster 2**. Contained 3,681 concepts (386 unique) and 3,273 patients who were 96.3% control (31.5% of all control patients). *Top-three concepts*: hypertrophy of tonsils and adenoids (31598; n=186), dyssomnia (435657; n=173), and knee pain (4150062; n=171). *Top-three unique concepts*: pH of urine by test strip (3022621; n=11), corneal foreign body (432768; n=7), and superficial injury of the hand (4086197; n=6).
- **Cluster 3**. Contained 6,715 concepts (2,177 unique) and 2,506 patients who were 27.1% CF (81.1% of all CF patients), 13.5% SCD (41.6% of all SCD patients), 9.9% CAH (32.8% of all CAH patients), and 3.3% PKU (35.3% of all PKU patients). *Top-three concepts*: protein [mass/volume] in serum or plasma (3020630; n=150), alkaline phosphatase [enzymatic activity/volume] in serum or plasma (3035995; n=125), and aspartate aminotransferase [enzymatic activity/volume] in serum or plasma (3013721; n=95). *Top-three unique concepts*: pancrelipase (919528; n=34), ciprofloxacin (19075379; n=23), and ultrase (19132375; n=21).
- **Cluster 4**. Contained 3,438 concepts (331 unique) and 2,051 patients who were 96.0% control (19.7% of all control patients). *Top-three concepts*: body height (3023540; n=623), body weight (3013762; n=346), and oxygen saturation in arterial blood by pulse oximetry (40762499; n=334). *Top-three unique concepts*: cesarean delivery - delivered (437942; n=8), unilateral incomplete cleft palate with cleft lip (132439; n=5), carrier of hereditary factor viii deficiency disease (4120610; n=4).
- **Cluster 5**. Contained 4,191 concepts (407 unique) and 2,488 patients who were 82.2% control (20.5% of all control patients) and 12.7% SCD (38.9% of all SCD patients). *Top-three concepts*: Haemophilus influenzae b vaccine (46276401; n=126), Prevnar 13 (40173508; n=126), and child examination (4088016; n=124). *Top-three unique concepts*: systolic blood pressure--standing (3035856; n=84), diastolic blood pressure--standing (3019962; n=83), and eosinophils [#/volume] in blood (3013115; n=34).

## Discussion

In this project, we perform a comprehensive characterization of patient representations learned from EHR data. Extensive ablation studies illuminate the impact of different combinations of sampling windows and data types on rare disease classification. These experiments were conducted in order to answer two research questions:

### Question 1: What combinations of data type and sampling window create the best patient representations and does performance differ by disease group?

Classification results suggest that there is no single best combination of sampling window and data type that performs best for all rare disease groups examined. This is illustrated in Figure 1; when using all concepts, the classification performance differed widely within and between each rare disease as a function of data type. For example, PKU, CF, and CAH patient representations performed best when learned using all available concepts from all data types (i.e., all condition, drug exposure, and measurement concepts). CF and CAH patient representations also performed well using only all drug exposure concepts, which makes sense clinically since there are targeted treatments for these diseases. In contrast, SCD patient representations performed poorly when using all concepts and only drug exposure concepts. Similar patterns were observed when examining the plots produced when using only those concepts that occurred before or after the index date. Although preliminary, these results provide evidence that the best patient representation is likely to depend upon the use case. For example, if the goal of an analysis is to create the best possible representation of a patient, it would be important to examine all of the different views that the data element configurations provide, and select only those elements which best reflect each patient. This may mean that only a portion of all available data and data types are needed. Constructing performant patient representations requires the right combination of data elements, which can be enhanced and improved by the insight generated by data-driven characterizations.

### Question 2: How does data-driven characterization of patient representation impact the explainability of downstream tasks like clustering?

When examining the impact of sampling window independent of a specific data type, using all of the available concepts performed best. When examining the impact of the different data types independent of specific sampling windows, using only drug exposure concepts performed best when clustering only rare disease patients, and using concepts from all data types worked best when clustering rare disease and control patients combined. Finally, when examining the impact of sampling window on data type, using only drug exposure concepts, regardless of the sampling window, resulted in the best performance when clustering only rare disease patients. When clustering rare disease and control patients, conditions performed best when using all available, drug exposures performed best when only those concepts before the index date, and using data from all of the data types performed best when using only those concepts after the index date. These results are directly related to explainability, which is best illustrated when examining the top features from the best performing combination of data type and sampling window. When clustering only rare disease patients, the best performing result was found for representations that were trained using all available drug exposure concepts. Of the four clusters, two were nearly pure to a single disease type (i.e., Cluster 1 was 99.7% CF and Cluster 3 was 92.8% CAH). For Cluster 1, the most frequent and unique drug exposure concepts were dornase alfa and two different strengths of the drug Creon. Both of these drugs are commonly used to treat patients with CF^28^. For Cluster 3, all but one of the most frequent drug exposure concepts were different strengths of levothyroxine sodium, a drug that is frequently prescribed for hypothyroidism^29^. The remaining clusters were mixed. Cluster 2 was 65.6% SCD and 26.9% CAH and contained frequent and unique concepts for different types of vaccinations. SCD patients are at risk of numerous infections, and vaccinations are a recommended preventative measure^30^. However, for CAH patients, which constitute a much smaller portion of the patients in this cluster, receiving vaccinations is considered routine preventative care. The final cluster contained 82.4% of PKU patients in addition to a small portion of patients from each of the remaining rare disease groups. The most frequent concepts in this cluster included amoxicillin, ibuprofen, and albuterol - medications frequently prescribed to most children, irrespective of their rare disease status. However, when examining the frequent concepts unique to this cluster, the two most important concepts were different forms of Kuvan, which is a dietary supplement prescribed in PKU to help lower phenylalanine levels in blood^31^. Similar explanations can be derived when analyzing the most important features in the clusters for the best performing clustering of the rare disease and control patients. Examining the most frequent concepts in each cluster confirms what one might expect, but examining the unique concepts highlights the different perspectives produced as a function of the different parameters used to learn each patient representation. Combining the insight provided by our approach, including simple queries like these, with typical model explainability methods could drastically improve the transparency and usability of complex learning pipelines. This should be explored in future work.

### Limitations and Future Work

This work relied on small samples of rare disease patients from a single hospital, which we were unable to validate using manual chart review (for privacy reasons) or with genetic data. Expanding the analysis to include more common and complex disorders is an important step to validate the usefulness of this approach for diseases which are less well-defined or more difficult to identify computationally (e.g., asthma and depression). It will also be important to perform additional experiments that directly assess and compare patient representations methods to traditional rule-based approaches like eMERGE’s PheKB^32^ and the Observational Health Data Science Initiative’s Atlas (https://github.com/OHDSI/Atlas/wiki). While we examined different types of structured data, we did not include unstructured data or quantitative values from measurements (i.e., test result values) or medications (i.e., quantity or strength). Examining these data is likely to be vital for improving the precision of the resulting representations. Finally, only a simple representation learning algorithm was used, future work should examine the utility of more complex algorithms, like language models, which have recently shown great utility, especially in situations where data is limited^33^. More complex techniques for clustering, which are able to account for the class imbalance within each disease, should also be explored in addition to exploring a larger number of clusters. When applying these methods, analyses should go beyond identifying the best and worst performing metrics and specify which differences are statistically significant. Finally, although not within the scope of the current project, future work should also investigate and incorporate methods like FIDDLE, which automate the processing and transformation of structured EHR data into concept- and patient-level feature vectors^34^.

## Conclusion

Advancing computational phenotyping will require a shift away from relying solely on iterative and expert-derived or rule-based cohort identification using predefined code sets and complicated clinical logic. Patient representation learning methods may be a promising solution, but their adoption in clinical settings has been somewhat limited due to scalability and explainability challenges. We demonstrated that the data type and sampling window directly impact classification and clustering performance, and these results differ by disease group. Although preliminary, these findings exemplify the importance of and need for data-driven characterization in patient representation-based CP development pipelines.

## Data Availability

Patient data produced in the present study are not available online.

